# Strength of attention-sampling parietal EEG theta rhythm is linked to impaired inhibition in adult ADHD

**DOI:** 10.1101/2020.06.03.20120964

**Authors:** Benjamin Ultan Cowley, Kristiina Juurmaa, Jussi Palomäki

## Abstract

Attention-deficit hyperactivity disorder (ADHD) in adults is understudied, especially regarding neural mechanisms such as oscillatory control of attention sampling. We report an EEG study of such cortical oscillations, in ADHD-diagnosed adults taking a continuous performance test that measures the ability to sustain attention and inhibit impulsivity for a prolonged period of time.

We recorded 53 adults (28f, 25m, aged 18-60), and 18 matched healthy controls, using 128-channel EEG. We analysed features with established links to neural correlates of attention: event-related (de)synchronisation (ERS/D), alpha and theta frequency band activation, phase-locking value (PLV), and timing-sensitivity indices; in frontal and parietal scalp regions.

Test performance distinguished healthy controls from ADHD adults. The ADHD group manifested significantly less parietal pre-stimulus 8Hz theta ERS during correct inhibition trials, less frontal & parietal post-stimulus 4Hz theta ERS during inhibition & response trials, and increased frontal & parietal pre-stimulus alpha ERS during inhibition & response. They showed significantly reduced fronto-parietal connectivity that lagged across trials and was strongly lateralised. In addition, they had reduced sensitivity to targets in stimulus-locking measures.

Building on the hypothesis of parietal attention sampling, our results suggest that ADHD adults have impaired attention sampling in relational categorisation tasks.

## Introduction

Attention-deficit hyperactivity disorder (ADHD) is among the most common childhood psychiatric disorders. ADHD is marked by persistent, age-inappropriate levels of inattention, hyperactivity and/or impulsivity. While the prevalence of ADHD declines with age, the condition persists into adulthood in approximately one third of those diagnosed as a child (1). There is thus a need for more research to study how the disorder manifests in adults. Here, we investigate how the neural correlates of attention differ between adults diagnosed with ADHD and healthy controls.

Both the clinical and scientific pictures of ADHD remain complex and multidimensional (2). Clinical diagnosis of ADHD operates at the level of heterogenous behavioural symptoms, subjectively assessed (3–5), with three sub-types of ADHD listed in diagnostic manuals: predominantly inattentive (ADHD-I, or ADD), predominantly hyper-active/impulsive (ADHD-HI), and combined (ADHD-C). Although diagnosis does not include behavioural attention testing, there have been efforts to develop indicative behavioural tests for ADHD (6). As yet, there remains no one-size-fits-all test that reliably classifies ADHD. Because of this, it is important to search for neurocognitive mechanisms that explain aspects of the disorder - mutual information from multiple mechanisms (e.g. behavioural and neural) could improve treatment targeting efficacy (7).

ADHD is characterised by deficits in completing cognitive tasks that challenge self-regulation of attention over extended durations. Thus, it is relevant to examine the interplay between sustained attention and inhibition, especially via the underpinning neural processes, such as regulation of perceptual processing by (dorsal or ventral) attention networks (8). High time-resolution, magneto- and electro-encephalography (M/EEG) studies have been vital in understanding the neurophysiology of ADHD by eliciting the mechanisms of attention. For example, cortical oscillations (i.e. rhythmic patterns of neural activity) have long been suggested to play a role in communication between areas of the brain (9), and have been shown to be altered in ADHD subjects (10) by measurement of event-related synchronisation.

One influential model says that ADHD is associated with deficient inhibitory control, which affects other executive functions (EF) such as sustained attention and working memory (11). The deficits in attention, inhibition, and working memory suggest that activity in the frontal cortico-striatal network and frontoparietal attention network are altered in ADHD (12), and may be linked to aberrant default mode network (DMN) suppression (13), or alpha-band oscillation suppression (14). Both volumetric alterations and changes in task-dependent BOLD (blood-oxygen level dependent) signals in prefrontal-striatal circuits support this view (15). Mowinckel et al. (16) also found causal support for these ideas from fMRI functional connectivity analysis of a decision-making task in a methylphenidate intervention. They illustrated that aberrant within-network connectivity was alleviated by treatment, but reduced sustained DMN suppression was not – implying that adults with ADHD may continue to experience debilitating effects of increased neuroenergetic demand over longer periods (attention fatigue), even when medication helps to normalise short-term performance (as predicted by the ‘cognitive-energetic model’ of ADHD (17)).

Sustained attention may be coordinated by patterns of synchronous oscillatory neural activation (18) – which are therefore a compelling target of study through several related measurements, including power, phase, and synchronisation.

Decline in sustained performance has been associated with changes in the ratio of frontomedial theta (4-8 Hz) to occipital alpha (8-12 Hz) power (18). This could possibly be due to reduced attentional sampling (at theta frequencies) and DMN suppression (at alpha frequencies), driven by neuroenergetic fatigue (17). Errors due to attentional lapses are sometimes followed by decreases in alpha power (19), which might function to refocus attention on the task at hand and inhibit task irrelevant processes (18).

Moreover, increased frontomedial theta power has been linked to monitoring of task-relevant activities and cognitive control, both of which are implicated in sustained attention (18, 20, 21). Frontomedial theta increase may also reflect mental fatigue (22), which consequently challenges the ability to sustain attention. Frontomedial theta power tends to increase during sustained performance tasks (for example, Wascher et al. (22)’s 4h-duration spatial stimulus-response task), correlates with increases in commission errors (incorrect responses) and reaction times (RT), and predicts post-error RT slowing (18).

Klimesch (9) proposed that reduced alpha desynchronisation is related to attentional control and inhibition, and theta synchronisation to task-specific semantic processing. Alpha oscillations tend to reduce in amplitude (due to neural desynchronisation) during effortful cognitive processing; when event-driven, this is called event-related desynchronisation of alpha (ERD). In contrast, theta oscillation amplitudes tend to increase with increasing cognitive effort (23), i.e. event-related synchronisation of theta (ERS) (24, 25).

Oscillations can become entrained (phase-locked) to predictable stimulus onsets. That is, the oscillatory phase (that facilitates the processing of subsequent stimuli) becomes more likely to coincide with the expected stimuli (26). This entrainment can be observed already in the pre-stimulus period (10, 27). Calderone et al. (10) suggest studying selective attention as indicated by oscillatory entrainment in clinical conditions such as ADHD. In lab conditions people with ADHD tend to be ill-prepared for incoming stimuli that may require a response (28). Several studies have linked the entrainment of pre-stimulus alpha oscillations to fluctuations in visual awareness and sustained attention (29–35).

### Attention Testing

Continuous Performance Tests (CPTs) are developed to measure sustained attention and inhibitory control. Among CPTs, the Test of Variables of Attention (TOVA) is considered a gold standard to detect attention problems typical for ADHD. TOVA is used on all age groups, both clinically and academically. It tests inhibition and the ability to sustain attention, with a monotonous hybrid Go/NoGo target classification task (6, 36), where subjects must respond to targets and inhibit response to non-targets – see Figure 1. TOVA software performs normative classification of test subjects’ performance as “not within normal limits”, “borderline” or “normal”. Many studies have used TOVA to study ADHD-diagnosed individuals *behaviourally*, primarily among children (for recent work see (37, 38)). Far fewer studies have measured EEG activity during TOVA (39, 40), and of these the only one focused on adults in particular was examining substance use disorder, not ADHD.

**Fig. 1.**
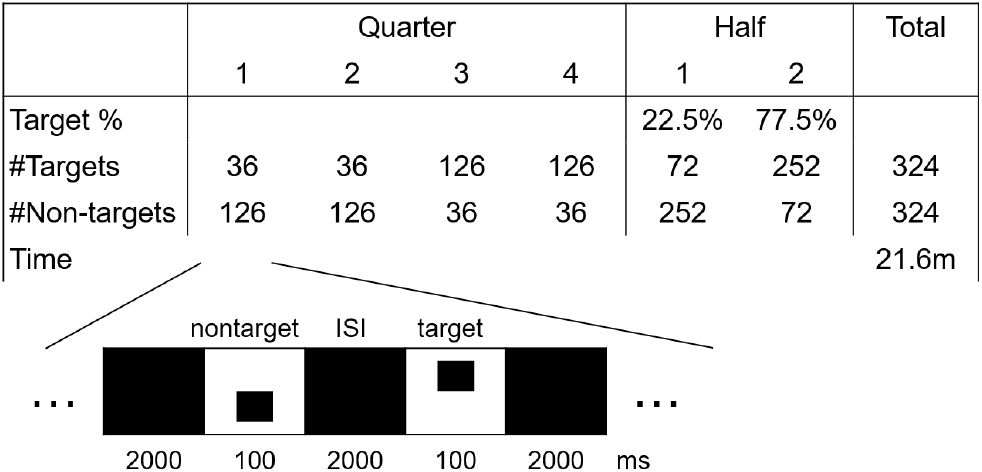
TOVA experiment protocol design. Above: number of targets and non-targets in each quarter and half, with target-to-nontarget frequency and total time. Below: individual trial structure, timings, and stimuli appearance.

Some previous studies have examined EEG activity during other CPTs while comparing individuals with ADHD to healthy controls. Loo et al. (41) found increased cortical arousal in the ADHD group during Conners’ CPT (42, 43), indicated by attenuated frontal and parietal alpha power. This effect increased towards the end of the task. However, they did not find any differences between groups in theta power.

### Research Questions

The aim of the present study is to examine the behavioural performance and cortical oscillations (measured with EEG) during TOVA, comparing adults with ADHD to a healthy control group. In particular, this study explores (a) how neural correlates of visual attention differ between groups throughout TOVA, and (b) how the increasing demands of sustained attention during TOVA affect oscillatory EEG and behavioural performance in adults with ADHD. We focus on the following research questions (RQs):

#### RQ1

How do ADHD and control groups differ in neural correlates of visual attention? We examine the temporal dynamics of frequencies linked to attentional regulation (particularly alpha and theta), in frontal and parietal regions-of-interest (ROIs), using event-related spectral perturbations (ERSP), event-related potentials (ERP), and phase-locking values (PLV).

#### RQ2

Does frontomedial theta power change during TOVA; is there a difference between the ADHD group and controls, or a group × time segment (first, second, or third half of the TOVA task) interaction?

#### RQ3

Does parietal or frontal alpha power change during TOVA; is there a difference between the ADHD group and controls, or a group × time segment interaction?

#### RQ4

Does pre-stimulus occipital alpha power change during TOVA; are there differences in pre-stimulus occipital alpha power between groups or between TOVA conditions (infrequent vs. frequent target mode), or a group × TOVA condition interaction?

## Methods

The measurement consisted of visual TOVA CPT with EEG recording, gathered as part of a larger project detailed in Cowley et al. (44). An ethical approval was obtained from the Ethical Committee of the Hospital District of Helsinki and Uusimaa. Each participant gave informed consent in accordance with the Declaration of Helsinki.

### Participants

We recruited 53 adults (25 males, age M=36.26, SD=10.22) diagnosed with either ADHD (n=44) or ADD (n=9) – we refer to both as ADHD group – and 18 adults (6 males, age M=32.78, SD=10.82) with no diagnosed neuro-cognitive deficits or ongoing medication for ADHD/ADD as a healthy control group. The healthy controls were significantly more likely to have TOVA performance within normal limits compared to ADHD adults (as detailed in the results section). All participants had normal or corrected-to-normal vision. The groups did not differ in terms of age, gender, or handedness. Due to reasons of data quality (detailed below) a number of participants were dropped from different EEG analysis stages. The final analysed number of participants ranged from 39 to 49 in the ADHD group, and from 14 to 18 in the control group. The control group size was always at least 33% of the size of the ADHD group, often closer to 40%, and thus the statistical power of our tests is not substantially affected by this size disparity.^1^

For the current study, inclusion criteria for the ADHD group were 1) pre-existing diagnosis of ADHD/ADD, 2) no neurological diagnoses, 3) age between 18-60, 4) scores on Adult ADHD Self Report Scale (ASRS) (45) and Brown ADHD scale (BADDS) (46) indicating the presence of ADHD, and 5) an IQ score of at least 80 using WAIS IV measured by a qualified psychologist (47). No strict cut-off values were used for ASRS and BADDS to indicate the presence of ADHD/ADD. Instead, exclusion was decided by the consulting psychiatrist, who conducted structured clinical interviews with participants as per the guidelines of the Diagnostic Interview for ADHD in Adults (DIVA 2.0, (48)). Comorbidities were evaluated during the clinical interview, and exclusion criteria included outlier scores in scores of Generalized Anxiety Disorder (49), Beck Depression Inventory (50), Alcohol Use Disorders Identification Test (51), the Mood Disorder Questionnaire (52), test of prodromal symptoms of psychosis (53), and the Dissociative Experiences Scale (54). The psychiatrist made a final assessment of the balance of symptoms contributing to patients’ presentation.

Within the ADHD group, there was a small difference in the ASRS hyperactivity-impulsivity scores depending on the diagnosis (*F*(1,50)=5.01, *p*<.030, *r*^2^=.09). As expected, participants with ADHD-HI or ADHD-C diagnoses had higher hyperactivity-impulsivity scores (M=6.33, SD=2.56; the groups were combined) than those with an ADHD-I diagnosis (without hyperactivity-impulsivity; M=4.22, SD=2.59). There was no significant difference in the ASRS inattention scores between groups, and most analyses thus treat all ADHD subjects as one group.

### Procedure

The behavioural and EEG data were measured in a 2.5–3 hour multi-task session, gathered in an electrically-shielded and sound-attenuated room. TOVA was administered towards the beginning of the measurement session (as suggested by Greenberg and Hughes (55, p-24)). The full session included preparation (30-40 min), pre-test baseline measurement (5 min), TOVA (22 min), resting state vigilance measurement (20 min), a novel CPT (30 mins; designed by (56)) and a post-test baseline measurement (2 mins).

Participants were asked to abstain from taking their ADHD medication for 48 hours prior to the EEG measurement (washout period). They were also advised not to take any other stimulants immediately prior to the measurement (e.g. coffee, cigarettes, energy drinks) and to arrive as well rested as possible to the measurement.

Karolinska sleepiness scale (KSS) (57) was employed to control for participants’ sleepiness levels, as sleepiness can affect both sustained attention and EEG measurement. Participants tended to report being alert (on the scale 1-9, ADHD: M=4.60, SD=1.38, controls: M=3.73, SD=1.39; these mean scores indicated 3 “Alert”, to 4 “Rather Alert”, and 5 “Neither alert nor sleepy”).

### TOVA

We administered TOVA visual version 8, consisting of the test software, USB relay hardware, a low-latency microswitch hardware response button, and Synchronization Interface hardware for test-to-EEG amplifier synchrony (all products of the TOVA Company).

TOVA presents a target and a non-target stimulus, shown in Figure 1. The participant’s task is to respond to targets by a button press of their dominant hand, and refrain from responding to non-targets; i.e. it has a Go/NoGo design. The participants are advised to respond as accurately and quickly as possible. A practice test of approximately 20 trials is administered before the actual test. Trials can be: correct responses, commission errors (incorrect responses), omission errors (incorrect inhibition) and correct inhibition.

TOVA has two consecutive conditions: infrequent and frequent target mode. During the first half (H1), the target stimulus appears infrequently (72 targets, and 252 non-targets, i.e. 22.5% target probability). During the second half (H2), these frequencies are reversed (252 targets and 72 non-targets, i.e. 77.5% target probability). Participants are not informed about this transition. Each stimulus is presented for 100 ms and separated from the next stimulus by a 2000 ms inter-stimulus interval (ISI). The anticipatory cutoff time from stimulus onset is 150 ms, meaning that responses given 0-150 ms after stimulus onset are considered invalid. The duration of TOVA is 21.6 minutes, each condition lasting 10.8 min.

### TOVA Variables

Analysis was performed on standardised TOVA scores, which are formed by comparing individual performance against a normed sample population (N=1596) (55). The comparison group for each participant is determined by their age group and gender.

Five standardised scores are available: 1) mean response time (RT) – mean of correct response times in milliseconds (ms); 2) response time variability (RTV) – standard deviation of mean correct RT; 3) commission errors – incorrect responses to a non-target stimulus; 4) omission errors – incorrect responses to a target stimulus; and 5) d’ (d prime) – ratio of hits (correct responses) to false alarms (commission errors). One participant who showed strong evidence of symptom exaggeration (Symptom Exaggeration Index, SEI) was excluded from further analyses.

For normative comparison, TOVA is advised to be measured between 6h00 – 13h00. Our group-average measurement times were close to or within the normed hours: ADHD M=13h45, SD=2h26 (35 after 13h00); control M=12h49, SD=3h02 (7 after 13h00), and the starting times did not statistically differ between the groups.

### EEG measurement and preprocessing

EEG was measured using Biosemi ActiveTwo equipment with 128 active electrodes, which is similar to the International 10-20 electrode placement system. Active electrode CMS (Common Mode Sense) and passive electrode DRL (Driven Right Leg) were used to create a feedback loop for amplifier reference. Electro-oculography (EOG) was recorded using bipolar montage: horizontal EOG electrodes were attached to the outer canthi of both eyes; vertical EOG electrodes were attached above and below the left eye. Electrode offsets (running average of voltage at each electrode) were kept below +/−25mV.

During the TOVA, participants were advised to relax and sit as still as possible, avoiding excessive movements, and to fixate the middle of the screen shown as a small white dot between stimulus presentations. Three participants were excluded due to technical problems with event codes in the EEG data. The final number of available participants and trials available for each statistical EEG analysis is summarised in the Appendix (Table A1).

The data was preprocessed using Computational Testing Automated Preprocessing toolbox (CTAP) (58, 59), based on EEGLAB (60) for Matlab. The data was re-referenced offine to the average of the two mastoids. The data was then low-pass filtered at 45 Hz and high-pass filtered at 2 Hz. CTAP was used to detect eye blinks using the method detailed in Cowley et al. (58). Validity of eye blinks detected by CTAP were examined for each participant by visual inspection.

Each participant’s continuous EEG and EOG data was decomposed using the FastICA algorithm (61). Independent Components (ICs) matching the CTAP-detected blinks were removed (58). Contaminated channels, epochs, and artifactual ICs were identified using FASTER (Fully Automated Statistical Thresholding for EEG artefact Rejection) methods (62). Data for automatically rejected channels was interpolated from adjacent electrodes. Fluctuations exceeding +/−80 microV in amplitude were discarded as bad segments. Any remaining ocular- or muscular-artifact ICs were detected and rejected manually by joint visual inspection of IC activations, scalp maps, power spectrum, and ERP image. On average 3% of channels and one IC per participant were rejected.

### Epoching for oscillatory power analyses

For event-related analyses, the EEG data were extracted into four types of epochs: commission errors, correct responses, omission errors and correct inhibition. Epochs lasted 2000 ms, with 1000 ms before and after stimulus onset. Epochs that contained a blink (as detected by CTAP) starting from 400ms before stimulus onset to 600ms after stimulus onset, were rejected. After this, too few error trials remained to conduct statistical analyses, thus analyses were run only on correct (inhibition and response) trials. Two participants (one in each group) were completely excluded to due high rate of epoch rejection. A baseline correction was applied to each epoch (from 1000 to 900 ms before stimulus onset) to counterbalance fluctuations in voltage offset across epochs and participants.

### Oscillatory power analyses

Logarithmic mean oscillatory power was calculated using Welch’s power spectral density estimation to calculate the mean log power spectrum (via EEGLAB’s spectopo function). Oscillatory power was calculated for each participant within three *time-segments* of the un-epoched data: during the first 5 minutes of TOVA (infrequent mode), during the middle 5 minutes (between infrequent and frequent mode), and during the last 5 minutes (frequent mode). Power was calculated for each 1 Hz frequency bin separately within the theta band (4–7 Hz) and within the extended alpha band (8–12 Hz).

For the theta band, power from electrodes in the frontomedial region (FCz, Fz, AFz, Fpz) were averaged together. For the alpha band, power from electrodes in the frontal region (F3, F4 and Fz) and the parietal region (P3, P4, Pz) were grouped and averaged within each region. In order to minimise the problem of multiple implicit comparisons, these electrode locations were chose a priori based on previous research on parieto-occipital alpha and frontomedial theta (18, 20, 21, 41). These calculations of alpha power follow the method of Loo et al. (41).

To examine the effect of TOVA condition on pre-stimulus activity, alpha power was also calculated for the 500 ms pre-stimulus period preceding correct trials for the two TOVA conditions, H1 and H2. Among all correct trials (both correct inhibition and correct response) 102 trials were randomly chosen among H1 and H2 separately for each participant. Some participants were excluded for these specific analyses due to a low number of usable trials (see Table A1).

Within these correct trials, pre-stimulus power was calculated for each 1 Hz frequency bin separately within the extended alpha band (8–12 Hz). Power from electrodes in the parieto-occipital region (P3, P4, Pz, POz, Oz, O1, O2) was averaged together. The electrode locations were chosen *a priori* based on previous research (31, 32, 35, 63).

### Event-related Spectral Perturbation calculation

To examine event-related oscillatory power dynamics for correct inhibition and response trials, event-related spectral perturbations (ERSP) (64) were calculated within the frontal and parieto-occipital ROIs defined above. Given that ROI-based ERSPs are not supported by EEGLAB, we created custom Matlab code to calculate ERSP matrices for each electrode individually and obtain the reported spectral power estimate as the mean of these^2^.

Spectral power was calculated in 54 frequencies from 4 to 30 Hz, in 200 time points (−583 to 581 ms), using Morlet wavelets with cycles increasing linearly from 3 to 11.25 in windows of 427 samples. Epochs were corrected by the joint group baseline. Note, the design of TOVA trials constrained the ERSP calculation to start at 4Hz, since lower frequencies returned very narrow windows.

The number of trials were balanced to ensure that any observed effect would be due to altered neural processing, not merely due to possible differences in behavioural performance (e.g. amount of correct response trials). For each participant, 108 correct inhibition trials and 108 correct response trials were randomly selected. Some participants were excluded from the ERSP analysis at this point, due to a low number of usable trials.

### Response-aligned ERP calculation

Response-aligned ERP images within frontal and parieto-occipital ROIs were generated by selecting 108 correct response trials randomly from each participant in ADHD and control groups. All trials were sorted by the corresponding RT value. A moving-window smoothing was applied: the control group had 1512 trials and window width 100; the ADHD group had more trials (4320) and a wider window of 285. Both target-locked and response-locked ERP images were computed. The corresponding ERP waves were computed as the grand-average of all electrodes in the ROI.

### Phase-locking value calculation

Phase-locking value (PLV) (65) for the upper theta band (6–10 Hz) was calculated for every pair of electrodes from frontal and parieto-occipital ROIs, both for whole trials and also, in order to examine temporal dynamics of PLV, in five 200ms windows from −200 to 800ms relative to stimulus onset. To obtain the PLV, data segments were padded with 300 samples, then bandpass filtered from 6–10 Hz with a finite-impulse response (FIR) filter of order 300. PLV was then calculated as per equation 1, where *N* is the number of trials and *θ*(*t, n*) is the difference between the instantaneous phase (computed using Hilbert transform) of the two signals at time *t* and trial *n*. The PLV magnitude is represented by color scales in Figure 4 below.

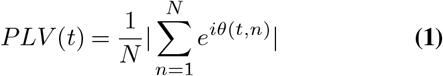

### Statistical analyses

#### Behavioural performance

Behavioural performance was analysed using a 2 x 2 two-way mixed design MANOVA, with Group (ADHD vs. control) as a between subjects factor and TOVA condition (infrequent vs. frequent) as a within-subjects factor. The DVs were mean RT, RTV, commission errors, omission errors and d’ (means and standard deviations of these variables are presented in the Appendix, Table A2). These were followed up by further DV-specific analyses, which are detailed in the Results-section. We also assessed how well behavioral performance in TOVA differentiates between ADHD diagnosed adults and healthy controls.

Effect sizes for repeated measures were computed as partial eta squared. Multiple comparisons were adjusted with Bonferroni correction, keeping the alpha at .05. Possible extreme outlier values were assessed by examination of studentized residuals for values greater than +/−3. We do note, however, that excluding outlier scores in a clinical sample is not straightforward, as high behavioural variability itself has been reported as a clinical characteristic in adults with ADHD (3). In total four outlier scores were excluded from the analysis of standardised scores. Excluding them did not affect the interpretation of results, while including them would have violated the assumption of homogeneity of variance-covariance matrices. When necessary, variables with skewed distributions were log-transformed. However, group means and standard deviations are reported in original untransformed format, as these are more meaningful to interpret. The assumptions of repeated measures ANOVA were satisfied on transformed data without outliers.

#### EEG data

Statistical analyses of EEG were run in Matlab (version 8.6), utilising EEGLAB toolbox (60) and in SPSS (version 21). Inspections of whether the data met the linear model assumptions (66) revealed moderate positive skewness and departures from normality (by Shapiro-Wilk) of alpha power. These were possible to correct by square root transformation; however the subsequent analyses produced very similar results for both original (logarithm-transformed mean power) and square root-transformed data. Therefore the results of analyses on original, logarithmic power are reported. Frontomedial theta power did not show departures from normality or skewness. Mauchly’s test indicated that the assumption of sphericity had been violated for all frequencies (*p*<.0005). Degrees of freedom were corrected using Greenhouse-Geisser estimates of sphericity to counteract the inflation of Type I errors.

The group difference of ERSPs was tested for statistical significance using permutation tests (60) (again based on custom code to handle ROIs). Significance was computed at two levels, *α*=0.05 (200 permutations) and *α*=0.0005 (2000 permutations), to illustrate a robust test statistic.

Group differences in correct-response trial ERP waves were tested using 2-sample Kolmogorov-Smirnov test of the mean amplitude within two time windows per trial: 150–250 & 330–430ms in target-locked, and −170–−70 & 0–100ms in response-locked trials (see Figure 4). These windows were centered on the N2 and P3 waves observed in the data. False Discovery Rate (FDR) correction (using the procedure described by Storey (67)) was used to correct for multiple comparisons at the *α*=0.05 level.

Difference between groups for PLVs was tested using boot-strapped 95% confidence intervals (CIs), obtained by computing PLV from 1000 resamples of the trials. Difference within trials (or 200ms sub-segments) was characterised by the *proportion of trial duration* for which a group’s CI was non-overlapping with the other group’s CI, e.g. Control group CI lower bound greater than ADHD group CI upper bound. This proportion value is used in Figure 4 (panel B) below to define line thickness in scalp maps.

Three 2×3 mixed ANOVAs were used to determine whether there were group × time-segment interactions in: frontomedial theta power, and alpha power in parietal or frontal regions. Also, a 2 × 2 mixed ANOVA was calculated to examine if there was a group × TOVA condition (frequent vs. infrequent) interaction in parieto-occipital pre-stimulus alpha power on correct trials.

## Results

We will first describe results from our behavioural analyses, and then focus on EEG results separately for each research question (RQ); we present exploratory results when they arise.

### Behavioural performance

Normed TOVA performance strongly distinguished diagnostic groups. Healthy controls were significantly more likely to have TOVA performance within normal limits compared to ADHD adults, as revealed by Pearson Chi-square (exact two-sided) test of independence (*χ*^2^(1)=7.66, *p*<.007).

In group-wise behavioural analysis (MANOVA), there was a significant TOVA condition × Group interaction (*F*(5,61)=2.63, *p*<.032, *η*^2^=.177). That is, the impact that TOVA condition (H1 and H2) had on performance (as measured by the linear combination of mean RT, RT variability, commission errors, omission errors and d’) depended on whether participants belonged to the ADHD group or controls. Thus, we followed up by examining the simple main effects of Group and TOVA condition. Behavioural results are illustrated in Figure 2.

**Fig. 2.**
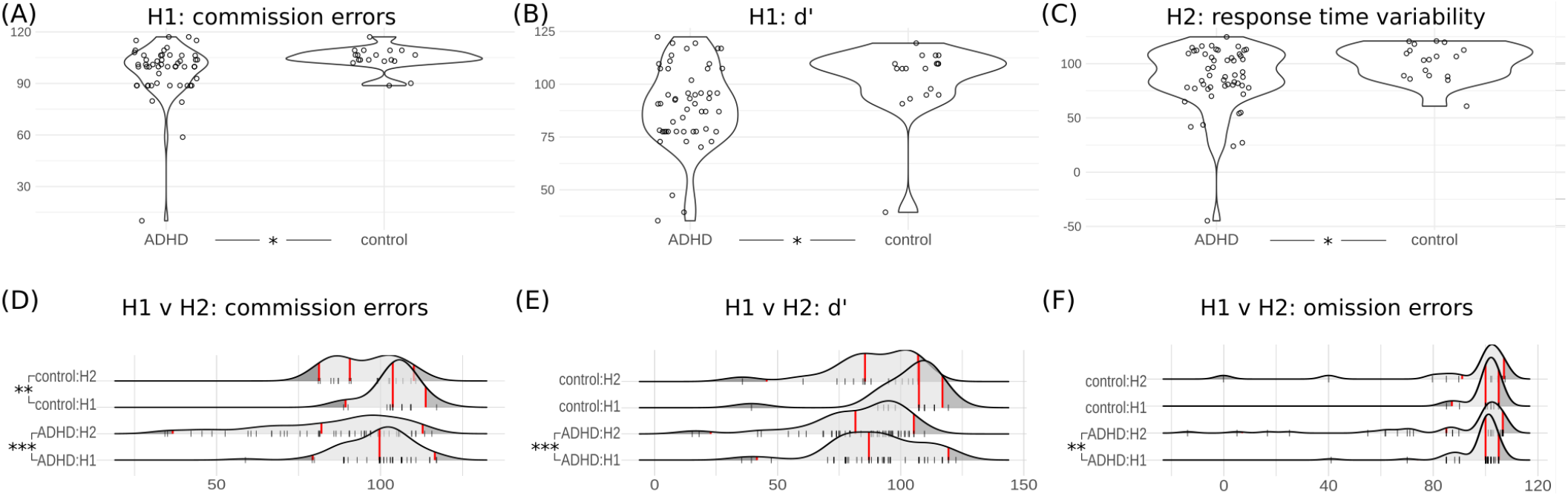
Behavioural results. **Panel A-B**: standardised scores for commission errors and d’, from TOVA first half. **Panel C**: response time variability standardised scores from TOVA second half. (Points in panels A-C shown with horizontal jitter for visibility). **Panel D-F**: rug-and-density plots show standardised scores for commission errors, d’, omission errors, across both groups and TOVA halves. Red vertical lines show the data mean, 2.5%, and 97.5% quantiles. *p* < 0.05 *, 0.01 **, 0.001 ***

First, we evaluated the difference between ADHD individuals and controls at the two TOVA conditions (H1 and H2) separately, by running two one-way MANOVAs for H1 and H2. The simple main effect of Group was not significant in either of these models (*F*s(5, 62) < 1.5, *p*s > .2). We then ran univariate one-way ANOVAs separately for all DVs instead of using a multivariate approach, again separately for H1 and H2. Within H1, there was a significant between groups difference (Bonferroni-adjusted for multiple comparisons) in commission errors (*F*(1,67)=4.12, *p*<.046, *η*^2^=.059) and in d’ (*F*(1,68)=5.30, *p*<.024, *η*^2^=.074): the ADHD group tended to produce more commission errors and had a worse d’ (the ability to discriminate between targets and non-targets) than the control group. Within H2, the ADHD group had significantly greater RT variability (*F*(1,68)=4.52, *p*<.037, *η*^2^=.064). See Table A3 in the supplementary materials for further details.

Next, we evaluated the difference between H1 and H2 (the difference in performance between the two consecutive conditions) separately for the ADHD- and control groups, by running two repeated measures MANOVAs. There was a significant simple main effect of TOVA condition in both the ADHD (*F*(5,44)=8.96, *p*<.0005, *η*^2^=.505) and control group (*F*(5,13)=7.63, *p*<.002, *η*^2^=.746): both groups performed worse towards the second half of TOVA. As before, we ran univariate repeated measures ANOVAs for each DV separately. Within the ADHD group, we found a significant effect of TOVA condition on commission errors (*F*(1,48)=26.08, *p*<.0005, *η*^2^=.352), omission errors (*F*(1,48)=11.15, *p*<.002, *η*^2^=.189) and d’ (*F*(1,48)=20.16, *p*<.0005, *η*^2^=.296): within the ADHD group performance deteriorated during the second TOVA condition across all measures. Within the control group, however, there was a significant effect of TOVA condition only on commission errors (*F*(1,17)=11.58, *p*<.003, *η*^2^=.405), and d’ (*F*(1,17)=8.67, *p*<.009, *η*^2^=.338). See Table A4 in the supplementary materials for further details.

### EEG results

In this section we focus on between-group analysis of EEG during non-error TOVA trials, for: frontal/parietal ERSPs, PLVs, and response-locked ERPs (RQ 1); band power estimates for frontomedial theta and parieto-occipital alpha (RQs 2-4).

### Alpha, theta dynamics on correct trials (RQ 1)

Figure 3 shows parietal (panel A) and frontal (panel B) baseline-corrected ERSPs for correct response and inhibition trials, for both Control group in column 1 and ADHD in column 2. Column 3 (panels A and B) shows the between-groups differences, masked by permutation-based significance testing where grey is not significant (n.s.), lighter-toned blobs are significantly different at *p*<0.05, and full-colour blobs are different at *p*<0.0005 (all tests uncorrected). Colour in the third panel represents Control minus ADHD ERSP, thus blue tones indicate ERS for ADHD.

**Fig. 3.**
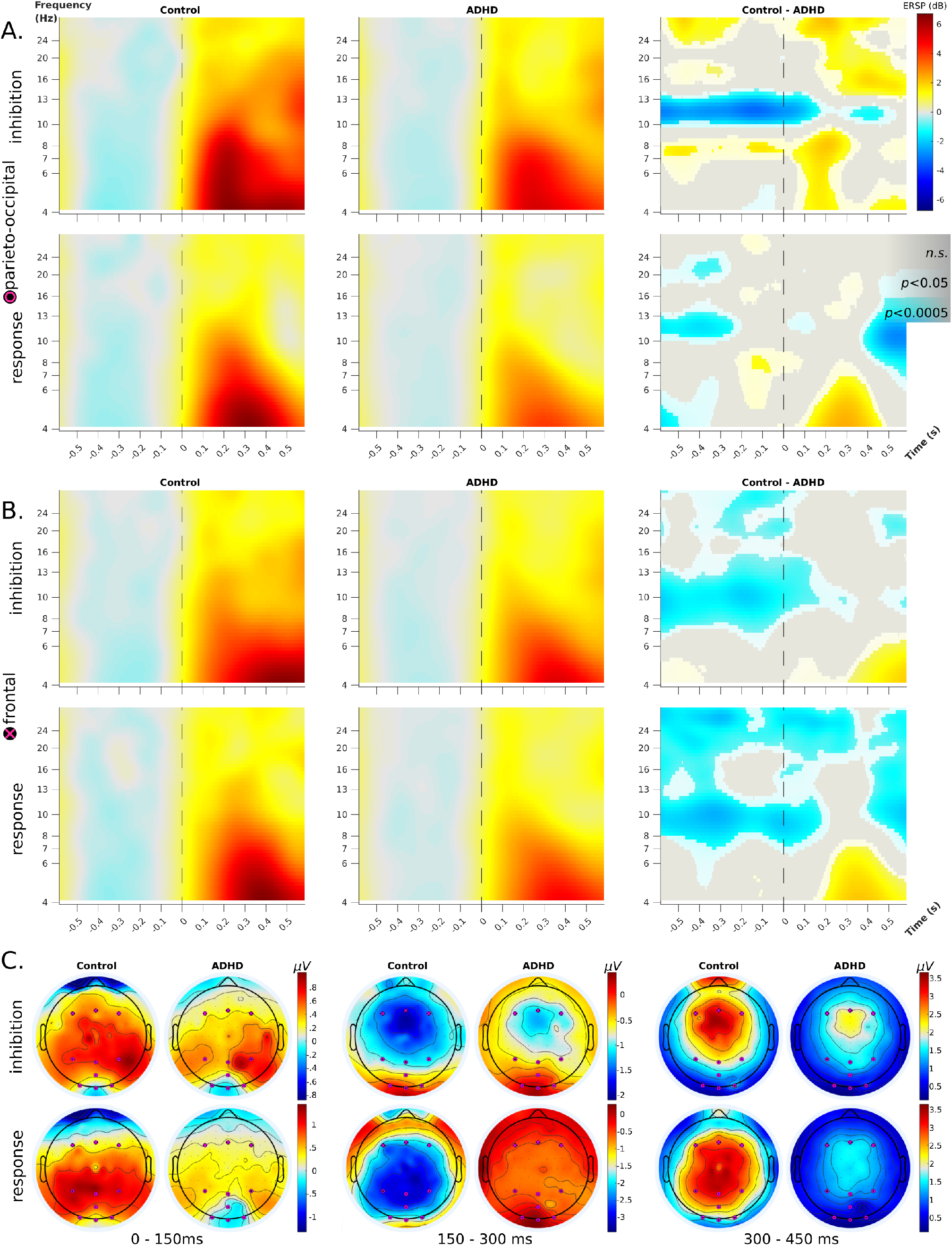
ERSP plots for correct inhibition and correct response trials, within parieto-occipital (panel A) and frontal (panel B) ROIs, with scalp maps (panel C). **Panels A, B**: ERSPs are locked to stimulus onset at time 0 (dashed lines); frequencies are plotted on a log scale. *Left* and *middle* columns show ERS/ERD for Control and ADHD groups, respectively, with removal of their shared mean baseline. The time-frequency data is averaged across all electrodes in the ROI; the scale of power perturbations goes from −6 to 6dB. *Right* column shows the log-mean difference between groups for the ROI-averaged time-frequency data; these plots are masked by a permutation-based significance test: grey is n.s., lighter-toned blobs are different at *p*<0.05, and full-colour blobs are different at *p*<0.0005 (all tests uncorrected). **Panel C**: whole-head scalp maps of amplitudes averaged within 150ms-long time windows across the post-stimulus duration of the ERSP plots. Colour scales are matched to the range of the data shared between groups within each condition and time-window. ROIs are shown as black circles with magenta symbols (frontal: x, parieto-occipital: o).

Main patterns of difference include: ADHD group has higher pre-stimulus alpha ERS centered on 10Hz frontally and 12Hz parietally, all conditions; Control group has higher 8Hz theta ERS in parietal inhibition trials; and Control group has post-stimulus bursts of 4Hz theta ERS across all conditions, varying slightly in timing. Other areas of difference do not show such consistent temporal patterns and are thus not considered further.

The between-groups differences in parietal theta arose ∼100ms after stimulus onset and continued at least for 300ms. The difference was about 2dB in magnitude, i.e. for Control group strongest theta ERS was 6dB and ADHD group was 4dB. The between-groups differences in frontal theta were slightly weaker and later.

The group-wise ERS (columns 1 & 2) indicate the pattern generating the differences: in all conditions the Control group shows a larger initial response at all theta frequencies, which is then sustained at 4Hz for inhibition trials. Parietal inhibition trials demonstrate the effect most clearly. Note, although these ERSP plots have lower bound at 4Hz, in supplementary material we include ERSPs calculated at 2 and 2.5 Hz which (despite their poor temporal resolution) show that the ERS is indeed focal at 4Hz.

Topographic scalp maps (Figure 3 panel C), computed in three 150ms time windows post-stimulus, illustrate the shift in location of maximum amplitude across time and between conditions/groups. Matching the ERSPs, the highest amplitude moves in the anterior direction over the trial, especially for inhibition trials. The main difference between groups, beyond the obvious difference in amplitudes, is that Controls have a slightly more posterior and left-lateralised focality of peak amplitudes.

### ERP results

Figure 4, panel A shows target-locked and response-locked ERP images (first two rows), plotted for both groups at both ROIs, along with the grand average ERP waves (third row). As expected (68), the ADHD group has more extreme RTs (particularly, very slow >500ms).

**Fig. 4.**
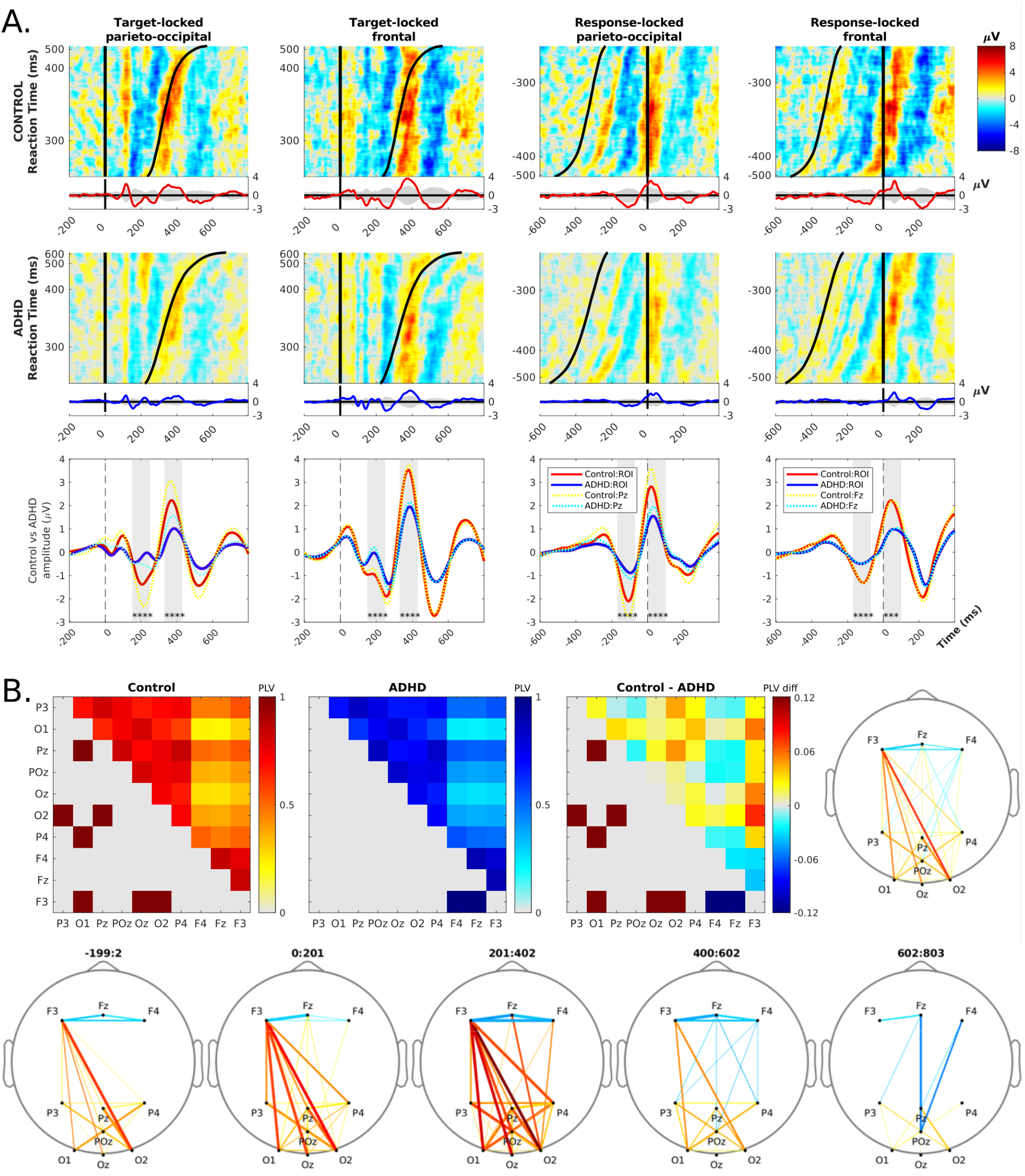
**Panel A**. Response-aligned ERP images at frontal and parieto-occipital ROIs during correct response trials. ERP images are matrices of activation data where each row is a single trial with colour-coded amplitude from −8 to 8 *µV* (cool and warm colors, respectively). On the left are target-locked trials from −200ms to +800ms of stimulus onset and time of response shown by the black sigmoidal curve. On the right are response locked trials from −600ms to +400ms of response time, and stimulus onset shown by the black sigmoidal curve. Cumulative ERP waves are shown below each ERP image. **First and second row** are Control and ADHD conditions: these ERP images show that stimulus-locked early waves (i.e. amplitudes at a fixed lag from 0), and the response-locked waves are both much stronger for control than ADHD group. Control group’s P3 also clearly begins *before* the response, in contrast to ADHD group. **Third row** are the ERP waves (solid lines) for each ROI, overlaid by the ERP (dashed lines) from each ROI’s central electrode (Fz, Pz). Vertical grey areas are test windows (aligned to N2, P3 in Target-locked trials) - group differences are highly significant. **Panel B**. Response-trial phase-locking values (PLV) between electrodes of our frontal and parieto-occipital ROIs, color-coded red for Control and blue for ADHD. **Top row**: in each matrix, upper triangle shows strength of PLV computed over whole trials for all-vs-all electrodes, while lower triangle shows connections where PLV was significantly greater than the other group for >30% of the trial duration. Third matrix, and scalp map, show the differences between group PLVs; scalp map line thickness shows proportion of trial duration where the difference was significant. **Bottom row**: similar scalp maps of the PLV difference in 200ms windows across the trial.

The control group has visibly stronger ERPs, in terms of both negative and positive power amplitudes. The maximum amplitude in the control group is *∼* 8*µV*, while in the ADHD group it is *∼* 4*µV*, and the grand average ERP difference tests are all highly significant (all had D≃0.1, *p* < 0.001).

The target-locked ERP images clearly illustrate a notable group difference: control participants experience much stronger phase-resetting in early waves (P1, N2) before 200ms. The response-locked ERP images show that Controls have a P3 preceding the RT by a substantial margin; in contrast, P3 in the ADHD group follows the RT.

### Phase analysis

We further investigated whether Control group’s superior performance reflected in stronger temporal coherence between target onset and neural responses, analysing phase relationships in correct response trials.

Figure 4, panel B further shows the contrast in phase dynamics with PLV for all-to-all ROI electrodes. Left & middle matrix (upper triangles) show within-group PLVs, while right matrix (upper triangle) shows group-difference of PLVs. Lower triangles of these matrices show the connections where PLV for one group was significantly greater than for the other group, measured by bootstrapped 95% CIs, for >30% of the trial duration. Local connections are typically strongest, but the notable exceptions are Control group’s F3– parieto-occipital connections.

The scalp maps do not apply the >30% threshold, but instead use this significance proportion to weight the line thickness: thus, only connections with long-lasting difference of 95% CIs are shown. The maps for 200ms segments (last row) illustrate the temporal dynamics for groups. Control begins *pre-stimulus* with greater PLV than ADHD between F3 and parieto-occipital electrodes (maximal for O2). During 400ms post-stimulus this difference strengthens and spreads across the whole fronto-parietal network (remaining strongly left-lateralised). In the last 400ms, the fronto-parietal connections grow stronger for ADHD than Control, but with opposite laterality: F4–POz and Fz–POz. Interestingly, for most of the trial ADHD group has much stronger PLV than Controls within the frontal ROI, until the last 200ms.

We also plotted phase-aligned ERPs and inter-trial coherence (for methods and figures see supplementary materials). Both these analyses focused on 10Hz, not only because it is the canonical value of peak alpha, but also the timing of TOVA stimuli is 10Hz, i.e. stimuli are shown for 100ms per trial.

Supplementary Figure 5/S1 shows ERP images at Pz, aligned to the pre-stimulus (−80ms) phase of 10Hz alpha, for randomly selected correct trials per-group and per-half. The figure confirms that the control group had much stronger phase resetting with higher amplitude early waves.

Supplementary Figure 6/S2 shows the 10Hz alpha inter-trial coherence (ITC) calculated per-group and per-half at Pz. Both group have significant 10Hz ITC peaking at around 200ms and 400ms, but it is markedly stronger for control group, by around 50%. For ADHD group, peak ITC droppped from H1 to H2, and the temporal extent of significant-ITC spread out, to begin earlier and end later. In contrast, for controls both H1 and H2 had almost equal profiles of significant ITC.

### Oscillatory power across time (RQs 2-4)

There was no significant group×time interaction, or a significant main effect of group in frontomedial theta power (RQ 2). There was however a significant main effect of time (*F*(8,58)=3.52, *p*<.002, *η*^2^=.327) on frontomedial theta in both groups. Pairwise comparisons revealed that only power at 7 Hz between time 1 and time 3 was significant (Bonferroni adjusted *p*<.05).

There was a significant main effect of time on alpha power (RQ 3), as measured from both frontal (*F*(10,56)=5.77, *p*<.0005, *η*^2^=.508) and parietal electrodes (*F*(10,56)=4.68, *p*<.0005, *η*^2^=.455). That is, power in the extended alpha band (8-12 Hz) increased with time in both groups. There was no significant group*×*time interaction in either electrode site.

In terms of parieto-occipital pre-stimulus alpha (RQ 4), there was no significant group ×time interaction, nor a significant main effect of group on correct trials. There was however a significant main effect of time (*F*(5,47)=3.4, *p*=.004, *η*^2^=302).

Tables of the complete set of results for the oscillatory band-power analyses and pairwise comparisons are presented in the supplementary materials (Tables A5 and A6).

### Sub-group differences

Since we did not find the expected between-group differences in oscillatory power, we split the ADHD group into *three* diagnostic groups for more detailed analyses: ADHD-HI/C, ADHD-I, and control.

There was a significant but small main effect of group on parietal alpha power (*F*(10,122)=2.35, *p*<.014, *η*^2^=.162) in addition to the significant effect of time segment (*F*(10,55)=5.82, *p*<.0005, *η*^2^=.514). Pairwise comparisons revealed that all time comparisons were significant (except time 2 vs time 3 at 8–9Hz, Table A5). The between groups difference lay between control and ADHD-I groups at 10-11 Hz (Bonferroni adjusted *p*<.05), wherein the ADHD-I group’s oscillatory power was twice as high (M=9.3, SD=6.2) as for the control group (M=4.5, SD=3.6). The effect was controlled for outliers defined as studentized residuals exceeding *±*3.

## Discussion

The present study explored cortical oscillations during sustained attention in adults with ADHD and in a healthy control group, measuring EEG while they performed the TOVA task.

### Group performance results

In terms of behavioural performance, both groups performed worse as the test progressed, from the low response-demand of the first TOVA condition (H1 – testing inattention, infrequent targets) to the higher response-demand of the second TOVA condition (H2 – testing inhibition, frequent targets). However the pattern of performance within and across test-halves differed between groups.

The ADHD group was affected by each TOVA condition in line with expected deficits of the diagnostic subtypes. During H1, testing inattention, the ADHD group had more commission errors and a lower d’ than the control group. During H2, testing inhibition, the ADHD group had more variable RTs than the control group.

Analysing performance between halves, both groups were affected by the change from H1 to H2, reflected in decreased d’ (possibly due to reduced vigilance) and increased commission errors (possibly due to fatigue). However, for the ADHD group the amount of omission errors also increased during H2, possibly due to mind wandering as suggested by the Cognitive-Energetic model of ADHD (17, 28, 69, 70).

### Research questions

We positively answered RQ1: Are there between-groups differences in neural correlates of visual attention? Indeed, both groups manifested ERS in the theta range on correct inhibition trials, but the amount of parietal ERS at 3-5 Hz was significantly higher for the control group. Parieto-occipital data showed Control had significantly higher ERS than ADHD at 8Hz, for inhibition trials from −500 to 200ms, and for response trials only around 100ms. The other consistent pattern was ADHD group’s significantly higher alpha ERS across the pre-stimulus period – this observation is in line with several prior studies that found deficient alpha *suppression* in ADHD during visual attention tasks (14).

Landau et al. (71) examined parietal theta in the context of visual discrimination-task performance, establishing a precise relationship between two-target spatial attention and gamma-band activity phase-modulated by a parietal 4 Hz source. More recently, Spyropoulos et al. (8) showed that (in macaque V4) parietal theta plays a role in modulating gamma-frequency coding of visual input, and in thus mediating visual attention. Evidence shows that sustained attention is a rhythmic sampling process occurring at a base rate of 8 hz when monitoring a single stimulus; this sampling rhythm decreases to 4 hz when there are two targets to monitor, and keeps decreasing with increasing number of targets (72). Observation of these effects in human electrophysiology indicates that they could be found also in scalp EEG, if generated strongly. TOVA sequentially presents two spatially-distinct (but otherwise identical) stimuli in a distraction-free context, thus providing ideal conditions to generate strong ∼4 Hz parietal theta ERS, exactly as observed. The sudden shift in Control group parieto-occipital inhibition trials, from 8Hz to 4Hz ERS, also supports this interpretation.

Further observations of parietal neural correlates of attention focused on the control group’s stronger phase-resetting in response to target stimuli. Controls had early ERP waves with significantly greater amplitude than the ADHD group (Figures 4, 5/S1); their P3 waves also coincided with responses, compared to ADHD which typically followed. Also, Control ITC profiles were both more focal and 50% stronger at peak (Figure 6/S2). Taken together, the evidence shows that ADHD participants were more weakly tuned to the periodic temporal profile of the TOVA trials, and thus had diminished capacity to predict the onset of the next stimulus (as expected from prior findings (28)). PLV results illustrated the group differences in connectivity – while Controls had stronger fronto-parietal and intra-parietal phase locking, the ADHD group had stronger intra-frontal PLV. And curiously, these patterns reversed towards the latter half of the trial. Also, when Controls had greater fronto-parietal PLV it was left-lateralised, in contrast to ADHDs who had right-lateralised greater fronto-parietal PLV.

Asymmetry was also reflected in the amplitude scalp maps in Figure 3, and indeed right-biased cortical asymmmetry in ADHD has been a common finding, for example in theta band among adults (73, 74). Hale et al. (73) suggested that “atypical rightward asymmetry should be broadly reflective of any form of non-optimized task-directed brain functioning”, which we elaborate on below.

Regarding the questions on band-power, RQs 2-4, results were mixed. RQ2 asked whether frontomedial theta power changed during TOVA, and our results were (surprisingly) negative, on balance. Frontomedial theta power did not increase during the increased demands on sustained attention in TOVA, with the exception of the increase at 7 Hz from first to last 5-minute time segment (see supplementary materials). No between-groups effect in frontomedial theta power was observed, in line with the results of Loo et al. (41).

RQ3 asked if alpha power at parietal or frontal locations changed throughout TOVA. Power in the extended alpha band (8-12 Hz) increased during TOVA in both groups, indicating decreased cortical arousal. Perhaps heightened demand on sustained attention resulted in fatigue and reduced alpha suppression capacity. This interpretation is in line with the finding that both groups performed worse during the second half of TOVA.

Again we found a null between-groups result; in particular, in contrast to Loo et al. (41), the ADHD group did not exhibit relatively lower parietal alpha power (more cortical arousal) compared with controls. Indeed, when we performed exploratory subgroup analysis, we found that the ADHD-I (predominantly inattentive type) subgroup’s parietal 10-11 Hz alpha power was twice that of the control group. This implies that these participants exerted less cognitive control and had diminished alpha suppression during TOVA, reflected as lower cortical arousal (i.e. higher alpha power). It is of note that differences were found only at 10-11Hz, precisely in time with the speed of presentation of TOVA stimuli.

Jang et al. (74) observed somewhat similar results, with weaker theta and stronger alpha for adults with ADHD traits (self-report only: not clinically screened), performing a spatial 2-back task. They also analysed PLV and found Controls had more significant connections in theta band in the first half of their trials. Although their sample does not allow for strong claims regarding ADHD, their converging evidence supports our results.

Finally, RQ4 asked if pre-stimulus occipital alpha power changed during TOVA, which was answered positively. On correct trials, pre-stimulus occipital alpha power increased towards the end of TOVA – potentially a reflection of reduced alpha suppression due to fatigue. However, this effect was not different between groups.

Overall, the deterioration of performance during TOVA appears a normal consequence of fatigue, brought on by paying sustained attention over an extended period of time. The lack of between-group differences in band power contrasts with the ERSP results, but the two analyses are concerned with different time scales. Taken together, the results of RQ1-4 indicate that in TOVA, our ADHD subjects were affected by trial-wise deficits, and not a deficiency of long-term neural energetics (17) – this might reflect that the task duration is well within the capabilities and self-efficacy expectations of adults, even with ADHD.

### Interpretation of results

*Synthesising all observed results*, we propose an interpretation based on deficient rhythmic attention-sampling in a cortical area responsible for relational processing, the posterior parietal cortex. This may be inferred from the nature of the TOVA task itself, as follows.

The TOVA task is temporally regular, meaning that every trial is predictable. For studying ADHD, this provides an advantage since the ADHD behavioural and neural responses can be compared to responses of a healthy control group which are facilitated by predictability. Thus, our particular observations point toward the ADHD behavioural deficit arising from reduced neural entrainment to task, possibly due to weaker endogenous attention sampling. The latter seems to be a parietal rather than frontal issue.

Specifically, based on the theory that attention continuously samples attended locations (at some fraction of an 8 Hz base rate (71)), our results suggest that it is the weaker parietal theta activation (reflected as reduced parietal theta ERS) that causes reduced phase resetting – i.e. reduced resources to monitor stimuli leads to weaker modulation of ongoing oscillations by those stimuli (or their predicted occurence). This consequently leads to weaker early ERPs (see Figure 5/S1) and smaller peak ITC (see Figure 6/S2). Thus, ADHD may have poorer TOVA performance because of weaker modulation of attentional sampling for stimuli requiring inhibition, reflected (for example) as significantly higher commission error rate in H1 (the half with frequent non-targets).

Further, TOVA is a relational classification task: Go/NoGo (respond/inhibit) to targets/non-targets, which share the same stimulus properties (smaller black rectangles within larger white squares), at two different spatial locations (above-below fixation). This relationship between target types permits a *unidimensional encoding*. Based on this property of the task, we can *hypothesise a cognitive mechanism* that could generate our above results, as follows.

Summerfield et al. (75) recently described a theory for the role of posterior parietal cortex (PPC, i.e. the dorsal visual stream, or the ‘what’ stream) in processing relational structure in the environment. In brief, Summerfield et al. (75) suggest that primate PPC supports learning and processing of relational structure, for example in visual scenes. They build on work establishing that PPC, in particular lateral intraparietal area (LIP, Brodman’s areas 39/40), provides spatially-selective coding for regions of ego-centric space that subserve functions including decision-making and top-down spatial attention (76). PPC neurons *also* code for abstract categories (77), in a scalar manner wherein unidimensional relations are coded asynchronously – if A is coded by many neurons, then B may be coded by relatively fewer. Since TOVA stimuli consist exactly of a unidimensional relational structure, every trial requires recruitment of the posterior parietal cortex to judge the presented relationship and trigger appropriate action.

The observed frontal theta ERSP difference, combined with the time course of PLV differences (Figure 4, panel B), suggests that an executive function deficit may also be at work. This is reflected in the timing of fronto-parietal synchrony, shown in later PLV windows (400-800ms) where ADHD Fz,F4–POz connectivity is ‘catching up’ compared to Control F3–parieto-occipital connectivity. The functional purpose could be compensatory, which reinforces the implication that the ADHD group was deficient in relational processing. This hypothesis refines the interpretation of the ERSP result: that not only does our ADHD group show reduced theta-rhythmic attention sampling, but they do so in the specific function of sampling relationships in the world.

### Limitations

The present study is among the first of its kind combining TOVA with an EEG measurement in adults (40) – and the first to focus on ADHD. As such, there were a number of limitations, both in the sense of constraints and issues.

#### Constraints

A number of aspects were suboptimal but were constrained by the fixed design of TOVA task, and size of sample locally available to meet the recruitment criteria.

For example, the small size of some of the TOVA norming groups, such as males aged 30-39 (n=4), might have increased the number of false positives. In the present study 19 participants (of which 15 with ADHD) belonged to this cohort. The low sample sensitivity and specificity highlight the issue of behavioural heterogeneity among people diagnosed with ADHD, and further motivates our sub-group analysis which found elevated parietal 10Hz alpha among the ADHDI group (compared to controls).

In terms of constraints of the task, TOVA does not aim to induce a large number of error trials, which limited the types of analyses available. For example, we could not subtract correct response trials from commission error trials, or *vice versa*. As a result, the ERSP plots for correct response trials (Figure 3) contain not only cognitive processing but also motor processing related to the response.

Finally, in relation to the oscillatory analyses of time-dependency (RQ2–4), it should be noted that the time-segment variable is confounded by the two TOVA conditions H1 and H2, the order of which are not counter-balanced in the TOVA software.

#### Issues

In order to recruit sufficient N the present study had a sample with a wide range of ages. There are well-documented age-related changes in tonic alpha and theta oscillations; for example, the age-related evolution of alpha and theta frequency spectra is nonlinear and may lead in opposing directions for alpha and for theta (9). However, given that our analyses were conducted within (not between) frequency bands, and the age range of ADHD and control groups were balanced, we do not believe this issue alters our interpretation of results.

We did not control for individual differences in alpha peak frequency (iAPF), that is, the single frequency in the alpha-band showing the highest power per individual. Counting iAPF as the central frequency of alpha, instead of 10 Hz, can be used to recalibrate all frequency bands and potentially account for clinical variation in EEG spectra (78). Individual differences in frequency spectrum may be more prominent than any age-related differences, particularly in the alpha and theta bands (9). On the other hand, for ADHD population such variation may be more prominent in children than adults.

The neuropathological heterogeneity of ADHD implies that distinct neurocognitive subtypes of ADHD may exist, and yet not map to symptomatic subgroups. Due to this, one criticism of ADHD studies has been that group-wise comparisons between ADHD participants and healthy controls are likely to yield small, hard-to-replicate effects (4). While the present study compared groups based on their diagnostic status and did find robust behavioural differences, also clustering participants as a function of both behavioural performance and cortical oscillations may find meaningful distinctions in test responses. This type of analysis-by-outcome (as opposed to analysis-by-diagnosis) might shed more light on how oscillatory dynamics are related to performance, and whether this relationship is altered in ADHD as compared to healthy controls. In the context of the present study, we did find strong differences between diagnostic groups, especially in event-related spectral and stimulus-locking analyses.

## Conclusion

In conclusion, this first-of-its-kind study examined the combination of TOVA CPT with EEG measurement for assessing neural correlates of ADHD in adults, an understudied question. We focused on participants’ diagnostic status and its relation to differences in oscillatory markers of sustained and relational attention during TOVA.

We showed that the ADHD group showed lower theta synchronisation during correct inhibition trials, and demonstrated reduced sensitivity to stimulus timing across several parietal measures. **The interpretation of these results is noteworthy, as it points to deficient attention sampling in relational categorisation tasks** (75). This result contributes to the overall understanding of the neuropathology of ADHD in adults.

Clinically, combining EEG measurement with TOVA has the potential to explain why differences in behavioural performance in TOVA arise. Identifying subgroups among ADHD diagnosed adults could eventually be used as an additional grouping variable to predict treatment outcomes.

## Data Availability

Participants did not explicitly consent to the release of the data recorded in the study, and thus datasets are not made publicly available.

## ACKNOWLEDGEMENTS

All authors co-wrote and approved the manuscript. KJ gathered and analysed the data, and created figures. BC conceived and designed the experiment, analysed data, and created figures. JP contributed to text and figures.

Authors wish to thank Mona Moisala and Marko Repo for data gathering; Otto Lappi, Alina Leminen and Eeva Palomäki for comments to the manuscript.

## Footnotes

1 See www.markhw.com/blog/control-size for a simulation analysis which suggests that a control group size 30% as large as test is comfortable

2 This preserves estimates of spectral power from separate electrodes, as opposed to averaging channels first and computing ERSP for the average, which may underestimate spectral power for electrodes at different phases.

