## Supplementary Material for "Strength of attention-sampling parietal EEG theta rhythm is linked to impaired inhibition in adult ADHD"

November 27, 2020

#### 1 Sample Specificity and Sensitivity of TOVA

Given the heterogeneity of ADHD and difficulty of its diagnosis especially in adults, we sought complementary evidence for our participants' attention deficit. In addition to self-report, we calculated the sample sensitivity and specificity of the TOVA test's ability to differentiate between participants with or without attentional problems. Note that, to get the two classes required for this analysis, the 'borderline' cases were coded as 'not within normal limits'.

TOVA company reports that their test has sensitivity and specificity of 0.8 (Greenberg, 2016, p. 21), giving an 80% chance that a person is correctly identified by TOVA as having (sensitivity) or not having (specificity) attentional problems typical for ADHD.

For our sample, there were 34 true positives (TP - expected 42 based on 0.8), 5 false positives (FP), 18 false negatives (FN) and 13 true negatives (TN - expected 14). Thus, for our sample, sensitivity was 0.65 and specificity was 0.72 – somewhat lower than expected.

#### 2 Supplementary ERSP and PLV

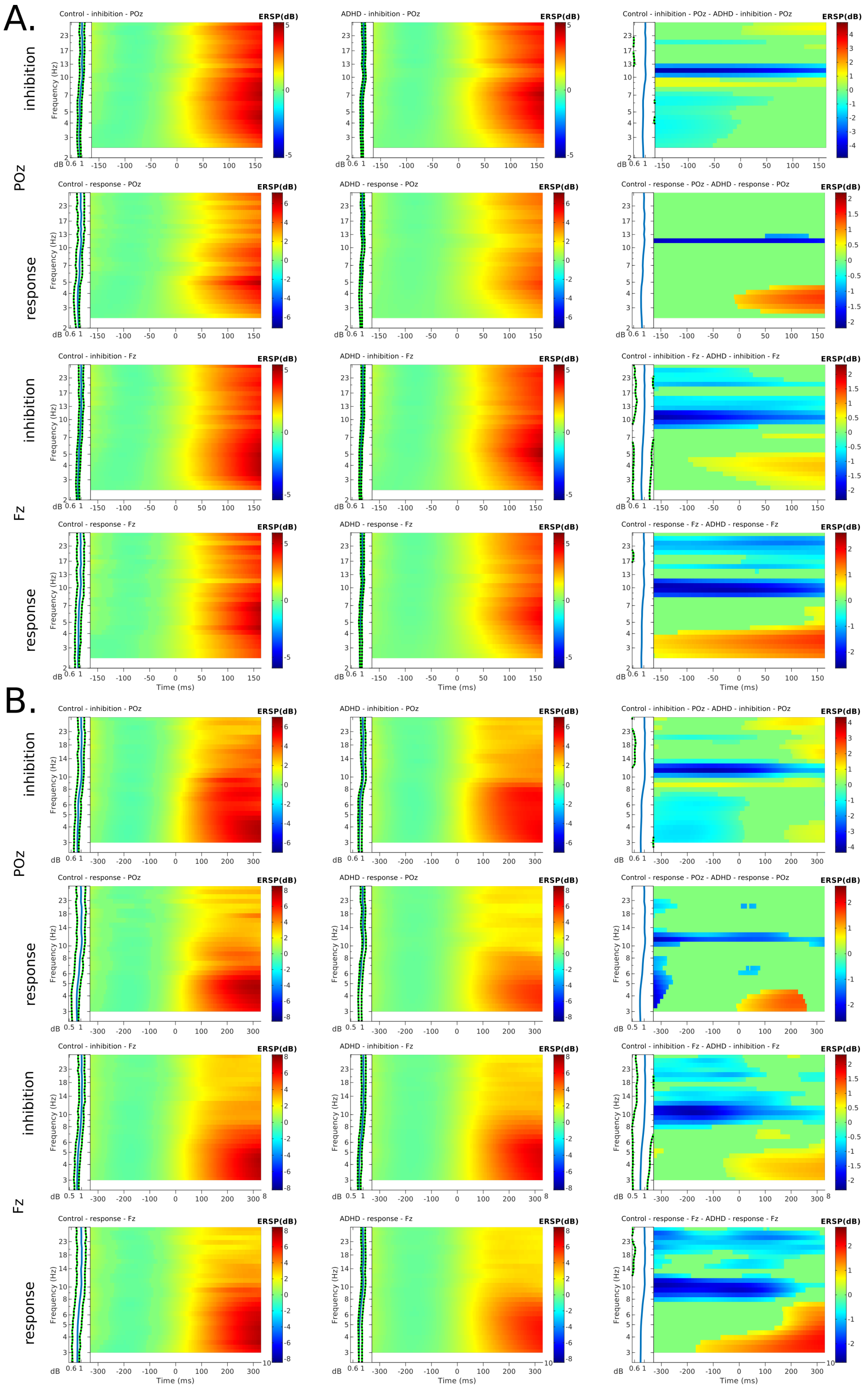

**Figure 5/S1.** Additional ERSPs which illustrate that the main post-stimulus ERS does indeed have a focus at 4Hz. These ERSPs have been calculated as those reported in main text, but for one electrode only (POz, Fz: the centres of the ROIs). Panel A shows ERSPs with lowest frequency 2.5 Hz, and Panel B shows lowest frequency 3Hz. As can be seen, the duration of the resulting windows is too short to see the most interesting temporal effects.

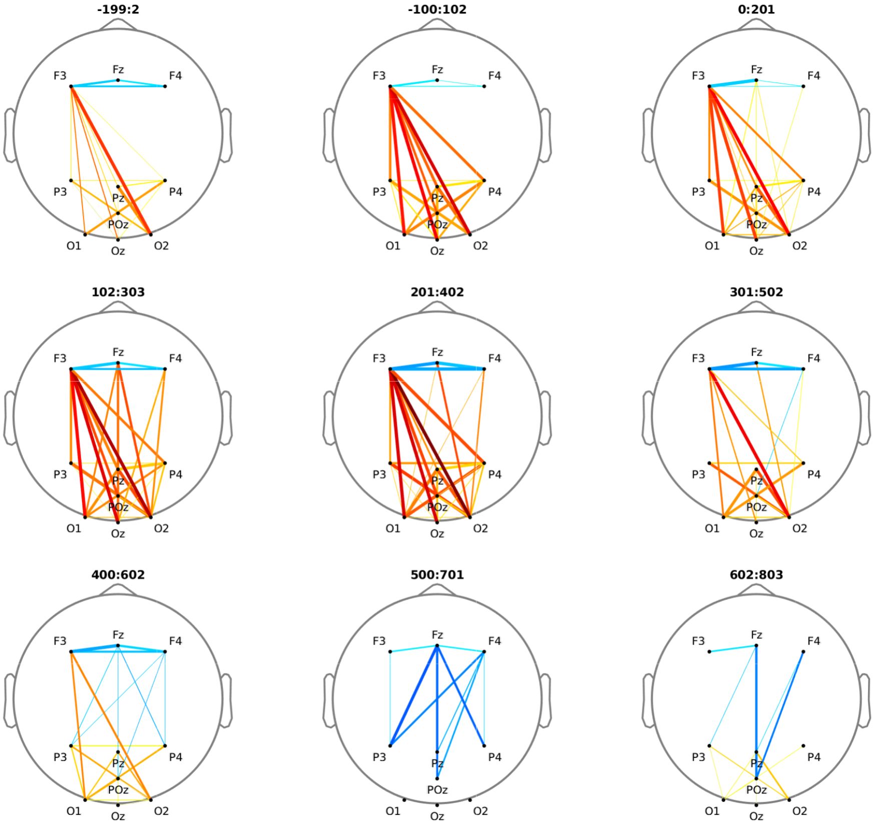


**Figure 6/S2.** This figure shows all nine of the PLV scalp maps generated by sliding windows of 200ms, overlapped by 50%. Thus, the PLV windows reported in the main text were selected from this set of nine, i.e. windows 1, 3, 5, 7, 9.

#### 3 Stimulus-locking analyses

**Phase-locking** As shown in Figure 3, we used EEGLAB to generate pre-stimulus alpha (10 Hz) phase-aligned event-related potentials images at electrode Pz, for randomly-selected correct inhibition or response trials, separately for the two TOVA conditions (H1 and H2). For the control group a moving-window smoothing of 100 trials was applied; and, as above, smoothing for ADHD was adjusted to 300 (H1) and 260 (H2). The center for phase alignment was set at 80 ms before stimulus onset.

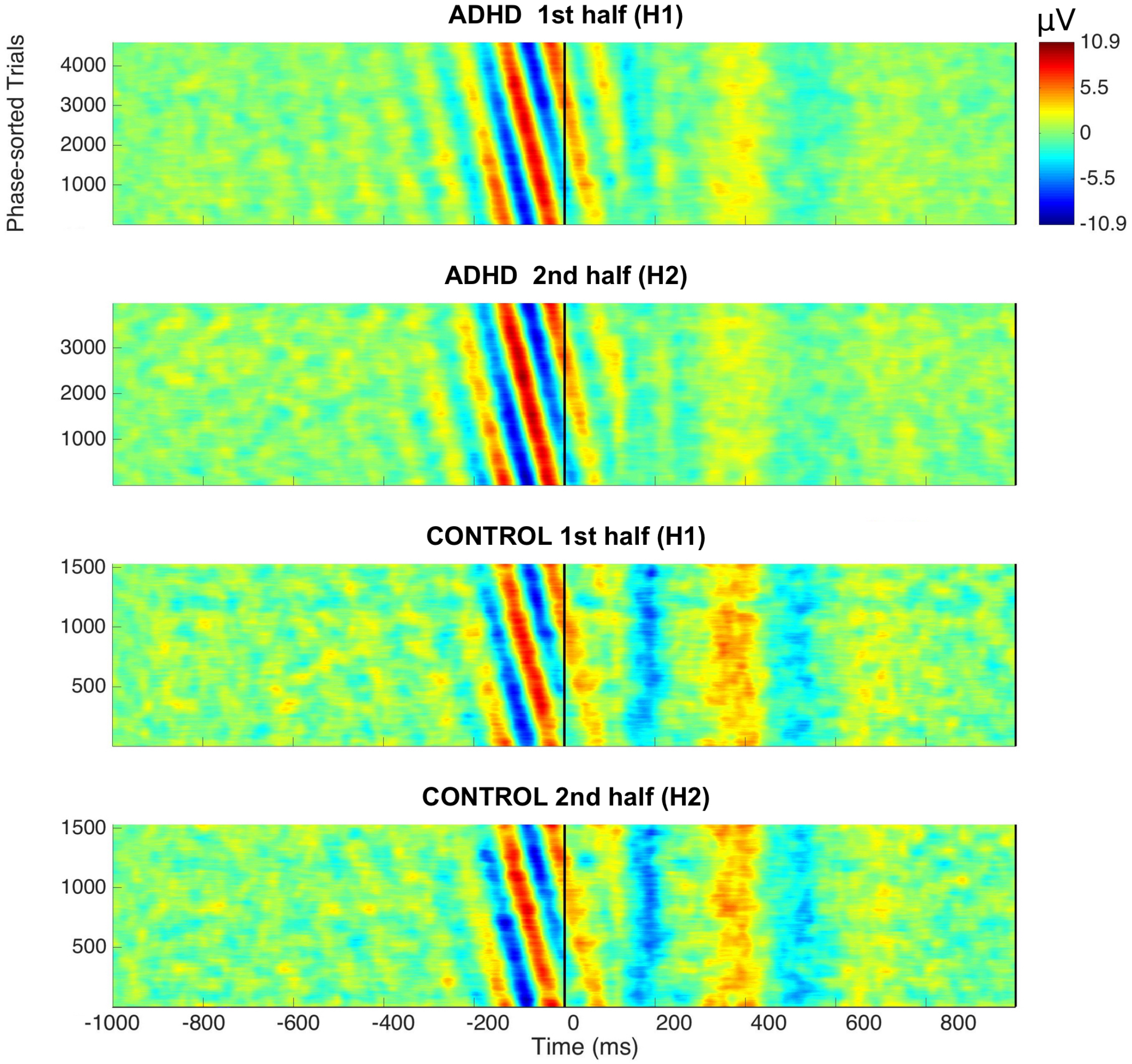


**Figure 7/S3.** Stacked correct-trial amplitudes, aligned to the pre-stimulus (-80 ms) alpha (10 Hz) phase, shown separately for the first and second halves of TOVA (H1 and H2) for both groups. Amplitude from -10.9 to 10.9 $\mu V$ is colour-coded to blue and red, respectively. The control group shows higher amplitudes in all stimulus-locked waves, i.e. phase-resetting reaction is enhanced compared to ADHD group.

**Inter-Trial Coherence** EEGLAB was used to compute 10 Hz alpha Inter-Trial Coherence (ITC) at Pz, separately for the two TOVA conditions (H1 and H2) and two groups, shown in Figure 3. EEGLAB computes statistical significance of the ITC via permutation testing of single-trial spectral estimates across latencies (Delorme & Makeig, 2004).

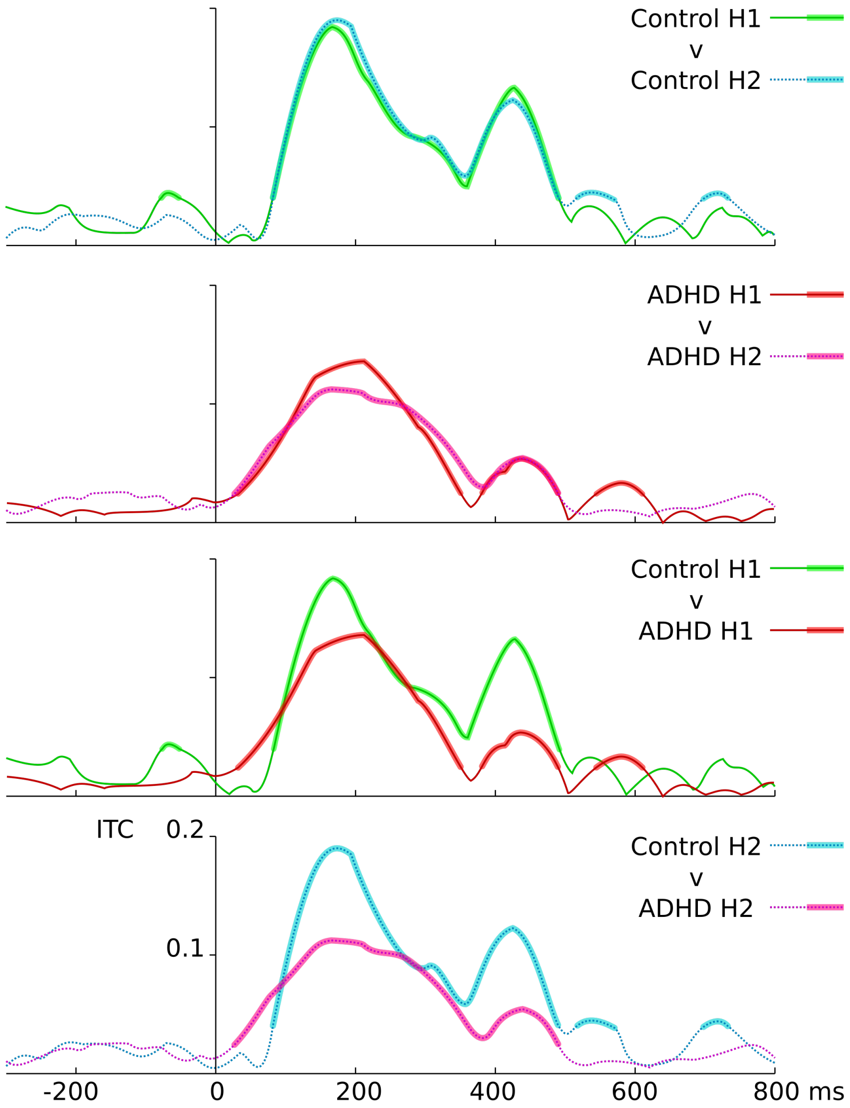


**Figure 8/S4.** Inter-trial coherence calculated for both groups and both TOVA conditions – all four ITC curves show peak around 200ms and smaller harmonic peaks, especially at 400. Wider lines show when ITC was significantly above chance level (the level of ITC which is significant depends on the sample, so no horizontal indicator is drawn).
**Top row**: control group condition H1 v H2 – almost no difference between conditions is seen.
**2nd row**: ADHD group condition H1 v H2 – a small reduction from H1 to H2 is seen.
**3rd row**: control H1 v ADHD H1 – substantial peak differences are seen at 200 (∼40%) & 400 (∼120%) ms. **4th row**: control H2 v ADHD H2 – large peak differences are seen at 200 (∼90%) & 400 (∼112%) ms. Control v ADHD comparisons also show that ADHD ITC is more dispersed, i.e. having weaker phase-locking to targets.

### 4 Descriptive Statistics and Analysis Results

**Table A1.** The final number of participants and trials per participant used for statistical EEG analyses, split by group.

| | TOVA condition or time segment | Number of participants | | EEG trials per participant |
|---|---|---|---|---|
| | | ADHD | Control | |
| Correct inhibition | H1–H2 | 42 | 15 | 108 |
| Correct response | H1–H2 | 40 | 14 | 108 |
| All correct trials | H1 | 45 | 15 | 102 |
| Continuous EEG | H2 | 39 | 15 | 102 |
| | segments 1-3 | 49 | 18 | – |

**Table A2.** Descriptive statistics of key standardised behavioural variables.

| | | ADHD | | | Control | | |
|---|---|---|---|---|---|---|---|
| Source | Time | Mean | SD | 95% CI | Mean | SD | 95% CI |
| RT variability | H1 | 89.02 | 28.32 | 80.88–97.15 | 100.19 | 17.57 | 91.45–108.92 |
| | H2 | 86.99 | 30.12 | 78.34–95.65 | 101.36 | 16.13 | 93.34–109.39 |
| Mean RT | H1 | 109.42 | 12.18 | 105.92–112.92 | 115.16 | 10.46 | 109.95–120.36 |
| | H2 | 109.04 | 14.82 | 104.78–113.30 | 111.21 | 9.70 | 106.38–116.02 |
| Commission errors | H1 | 98.90 | 10.83 | 95.79–102.01 | 104.56 | 6.58 | 101.29–107.83 |
| | H2 | 83.08 | 25.11 | 75.87–90.30 | 91.26 | 20.36 | 81.14–101.38 |
| Omission errors | H1 | 99.26 | 5.41 | 97.71–100.81 | 100.61 | 5.18 | 98.04–103.19 |
| | H2 | 75.95 | 55.29 | 60.07–91.83 | 90.70 | 27.68 | 76.94–104.47 |
| d’ | H1 | 92.18 | 16.88 | 87.33–97.03 | 102.20 | 17.68 | 93.41–110.99 |
| | H2 | 78.07 | 28.02 | 70.02–86.12 | 82.43 | 29.19 | 67.91–96.95 |

*Note.* Scores $>$ 85 are within or above normal limits, scores 80-85 indicate

borderline ADHD, scores $<$ 80 indicate performance that is not within normal limits.

**Table A3.** Main effect of group (MANOVA) and simple effects of group (separate ANOVAs) on each TOVA standardised variable, split by test halves.

| Source | Time | df | Error df | $F$ | $p$¡ | $\eta^2$ |
|---|---|---|---|---|---|---|
| Group (main effect) | H1 | 5 | 62 | 1.45 | .218 | .105 |
| | H2 | 5 | 62 | 1.26 | .294 | .092 |
| RT Variability | H1 | 1 | 68 | 2.71 | .104 | .039 |
| | H2 | 1 | 68 | 4.52 | .037* | .064 |
| Mean RT | H1 | 1 | 68 | 2.55 | .115 | .037 |
| | H2 | 1 | 67 | .055 | .815 | .001 |
| Commission errors | H1 | 1 | 67 | 4.12 | .046* | .059 |
| | H2 | 1 | 68 | 1.68 | .119 | .025 |
| Omission errors | H1 | 1 | 67 | .95 | .333 | .014 |
| | H2 | 1 | 67 | 1.18 | .282 | .017 |
| d' | H1 | 1 | 68 | 5.30 | .024* | .074 |
| | H2 | 1 | 68 | .68 | .412 | .010 |

*Note.* * significant at $p<.05$, Bonferroni adjusted for multiple comparisons.

**Table A4.** Simple effects of TOVA condition on standardised scores in each group.

| Group | Measure | df | Error df | $F$ | $p<$ | $\eta^2$ |
|---|---|---|---|---|---|---|
| ADHD | RT variability | 1 | 48 | .26 | .610 | .005 |
| | RT mean | 1 | 48 | .15 | .702 | .003 |
| | Commission errors | 1 | 48 | 26.08 | .0005*** | .352 |
| | Omission errors | 1 | 48 | 11.15 | .002** | .189 |
| | d' | 1 | 48 | 20.16 | .0005*** | .296 |
| Control | RT variability | 1 | 17 | .10 | .752 | .006 |
| | RT mean | 1 | 17 | 3.83 | .067 | .184 |
| | Commission errors | 1 | 17 | 11.58 | .003** | .405 |
| | Omission errors | 1 | 17 | 2.31 | .147 | .120 |
| | d' | 1 | 17 | 8.67 | .009* | .338 |

*Note.* * significant at p<.05, ** p<.005, *** p<.0005, Bonferroni adjusted for multiple comparisons.

**Table A5.** Effect of time segment on frontal (F3, F4, Fz) and parietal (P3, P4, Pz) alpha (8-12 Hz) power.

| Electrodes | Measure | df | Error df | $F$ | $p<$ | $\eta^2$ | Pairwise comparisons 1<2 | 1<3 | 2<3 |
|---|---|---|---|---|---|---|---|---|---|
| Frontal (F3, F4, Fz) | 8 Hz | 1.32 | 85.56 | 9.91 | .001** | .132 | ** | ** | ns |
| | 9 Hz | 1.36 | 88.54 | 22.05 | .0005*** | .253 | ** | *** | *** |
| | 10 Hz | 1.72 | 111.67 | 29.64 | .0005*** | .313 | *** | *** | *** |
| | 11 Hz | 1.71 | 111.38 | 38.30 | .0005*** | .371 | *** | *** | *** |
| | 12 Hz | 1.59 | 103.41 | 30.37 | .0005*** | .318 | *** | *** | *** |
| Parietal (P3, P4, Pz) | 8 Hz | 1.40 | 90.91 | 7.39 | .003** | .102 | * | * | ns |
| | 9 Hz | 1.50 | 96.58 | 18.14 | .0005*** | .218 | *** | *** | * |
| | 10 Hz | 1.50 | 97.49 | 23.20 | .0005*** | .263 | *** | *** | ** |
| | 11 Hz | 1.59 | 103.57 | 25.98 | .0005*** | .286 | *** | *** | ** |
| | 12 Hz | 1.52 | 98.48 | 27.94 | .0005*** | .301 | *** | *** | *** |

*Note.* * significant at p<.05, ** p<.005, *** p<.0005. Degrees of freedom are Greenhouse-Geisser adjusted for violations of sphericity. Time segments in the pairwise comparisons are *1* = first 5 minutes of TOVA (infrequent mode), *2* = middle 5 minutes (both infrequent and frequent mode), *3* = last 5 minutes (frequent mode).

**Table A6.** Effect of TOVA condition on parieto-occipital pre-stimulus alpha (8-12 Hz) power.

| Measure | df | Error df | $F$ | $p<$ | $\eta^2$ | Comparisons |
|---|---|---|---|---|---|---|
| 8 Hz | 1 | 51 | 2.96 | .092 | .055 | H1<H2 n.s. |
| 9 Hz | 1 | 51 | 4.28 | .044* | .077 | H1<H2* |
| 10 Hz | 1 | 51 | 9.27 | .004** | .154 | H1<H2** |
| 11 Hz | 1 | 51 | 8.08 | .006* | .137 | H1<H2* |
| 12 Hz | 1 | 51 | 13.27 | .001** | .206 | H1<H2** |

*Note.* Bonferroni adjusted for multiple comparisons: * significant at p<.05, ** p<.005, *** p<.0005.
